# Non-concussive Repetitive Head Impacts and Intense Physical Activity Elevate pT181-Tau and pT181/total-Tau in Plasma of Young Adults

**DOI:** 10.1101/2022.10.13.22281039

**Authors:** Martin Cente, Janka Perackova, Pavol Peracek, Marek Majdan, Igor Toth, Martin Mikulic, Jozef Hanes, Sara Porubska, Marian Spajdel, Barbora Kazickova, Igor Jurisica, Peter Filipcik

**Author notes:** Corresponding author Corresponding author: Assoc. prof. Peter Filipcik, PhD., Institute of Neuroimmunology, Slovak Academy of Sciences, Dubravska cesta 9, 845 10 Bratislava, Slovakia.

## Abstract

**IMPORTANCE:** Head impacts resulting in traumatic brain injury (TBI) lead to elevation of phosphorylated tau protein in plasma, the biomarkers for patients with acute and chronic TBI. Dynamics of phosphorylated tau protein level (pTau) and ratio of pTau and total tau in subjects after the non-concussive head impact was not yet investigated.

**OBJECTIVE:** To determine the acute effect of repetitive low-intensity head impacts on phosphorylated and total tau protein levels in plasma of young adults and to assess the effect of the head impacts on focused attention and cognitive flexibility.

**DESIGN:** The study cohort consisted of young professional soccer players performing intense physical activity without heading the ball (n=32) and physical activity including the heading (n=28).

**SETTING:** The effect of repetitive non-concussive head impacts was monitored in all athletes evaluating the parameters of physical activity, impact force during heading the ball, together with neuropsychological testing of cognitive attention and cognitive flexibility at multiple time-points before and during a set of controlled training exercises. The total and phosphorylated tau protein levels were determined using an ultrasensitive single molecule assay at 3 time points: before training, 1 h, and 24 h after each training session in a non-fasting state. The psychological scores were collected before and 1 h after the training using Trail Making Test.

**PARTICIPANTS:** The study included volunteer male college soccer players. The eligible participants were selected based on the similar demographic variables, such as age, years of performing organized sport, weekly sport activity and body mass index. The subjects with the history of previous TBI were excluded from the study.

**MAIN OUTCOMES AND MEASURES:** The primary study outcomes are the levels of total tau protein and tau protein phosphorylated at the Threonine 181 in plasma samples, and cognitive status of the study participants. The study hypothesis was formulated before the data collection.

**RESULTS:** We found significantly elevated levels of total tau and phospho-tau protein (pT181) in the plasma of soccer players one hour after physical exercise (Tau: 1.35-fold, 95% CI 1.2-1.5, *P* = .0001; pT181: 1.4-fold, 95% CI 1.3-1.5, *P* < .0001) and repetitive head impacts (Tau: 1.3-fold, 95% CI 1.2-1.4, *P* < .0001; pT181: 1.5-fold, 95% CI 1.4-1.7, *P* < .0001) in comparison to baseline. The pT181/Tau ratio was significantly higher 1 hour after the exercise and heading training, and remained elevated specifically in the heading group even after 24 hours (1.2-fold, 95% CI 1.1-1.3, *P* = .0024). Performance in cognitive tests revealed a significant decline in focused attention (TMT-A) and cognitive flexibility (TMT-B) after the physical exercise (TMT-A: 75.16 [95% CI, 68.3-82.01, *P* < .0001]; TMT-B: 80.26 [95% CI, 69.83-90.68, *P* < .0001]) and heading training (TMT-A: 33.76 [95% CI, 30.11-37.42, *P* = .0004]; TMT-B: 63.92 [95% CI, 56.99-70.84, *P* = .0434]). Nevertheless, the physical exercise of higher intensity without the heading had a more negative impact than heading training (TMT-A: *P* < .0001; TMT-B: *P* = .0207).

**CONCLUSIONS AND RELEVANCE:** Our study showed, that non-concussive repetitive head impacts elevate pT181-Tau and pT181/total-Tau in plasma of young adults. The increase of pT181-Tau was observed also after the intense physical activity alone, however the elevation of pT181/Tau ratio after 24 hours was highly significant and specific for non-concussive repetitive head impacts. The results support the idea that phosphorylated tau-enriched fraction of tau proteins may have long-lasting consequences in the brain of head-impacted individuals. The possible longitudinal disbalance of tau proteins induced by repetitive head impacts needs to be further investigated in larger and broader cohorts, and over longer period of time.

## INTRODUCTION

Neurodegeneration and dementia belong to the major public health, medical, and societal problems. The number of patients has been recently estimated at tens of millions of individuals. The Global Burden of Diseases study indicate that the number of people with dementia will almost triple in the coming decades, increasing from 57.4 million in 2019 to 152.8 million cases in 2050 ^1-3^, without disease-modifying therapy available. It is widely accepted that head impacts resulting in traumatic brain injury (TBI) are a significant risk factor for the development of sporadic tauopathies such as chronic traumatic encephalopathy (CTE), Alzheimer’s disease (AD), and other neurocognitive disorders ^4-6^. The mortality from neurodegenerative or neurocognitive disorders is about 3.5 times higher in professional soccer players compared to the general population, as documented in a large retrospective study ^7^. In American football players, the severity of CTE was found to be associated with the number of years played ^8^. However, for the consequences of low-intensity repetitive head impacts that are not followed by loss of consciousness (non-concussive head impacts), the data from the literature are inconsistent. Therefore, it is not clear whether or not such mild head impacts are associated with neuropathology ^9^, or what frequency of repeated head impacts may increase the risk of neurodegeneration.

Tau protein, a crucial microtubule-stabilizing protein plays an essential role in normal neuronal architecture and axonal transport. Disturbances in tau protein structure caused by posttranslational modifications such as phosphorylation, truncation, nitration, glycosylation, methylation and acetylation can be manifested in brain tissue as detrimental events leading to neurodegenerative diseases ^10,11^. It is not yet completely clear how the pathology starts at the molecular level; however, tau protein phosphorylated at specific sites (pTau) is considered a key pathogenic molecule that plays an important role in disease development and pTau is one of the diagnostic targets.

It was repeatedly demonstrated in experimental models that mild traumatic brain injury leads to tau protein hyperphosphorylation ^12-14^. Phosphorylated tau induces primary steps in generation of pair helical filaments and provokes the building of neurofibrillary tangles (NFTs), that block axonal transport which leads to the loss of intercellular connectivity and neuronal death ^15,16^. The accumulation of phosphorylated tau protein in perivascular and interneuronal space is another critical step in the development and spreading of neurofibrillary degeneration ^17-20^.

In spite of the indications from animal model studies, no causal link between head impact and tauopathy has been shown at the molecular level in humans. One of the reasons is the inability to obtain a definitive diagnosis of a specific tauopathy in humans without postmortem examination. Tauopathies therefore develop undetected before the onset of clinical signs and become apparent only after a long period of latency. During this period, NFTs composed of pTau deposit in specific regions of the brain, and lead to development of symptoms characteristic of different forms of neurodegenerative diseases, including CTE, AD, and others ^21,22^. Importantly, in patients at the risk, the correlations between imaging (tau and amyloid PET) and CSF or plasma tau protein levels are highly convincing ^23,24^.

The examination of peripheral plasma, which become available just recently, is much more convenient and widely accessible than CSF. Availability of novel hypersensitive tests allows detection of very small amounts of neuroproteins in plasma, which can potentially revolutionize our knowledge in the field ^25^. Quantification of tau proteins phosphorylated at specific residues in plasma may improve early clinical diagnostics of neurodegeneration ^26^. The pT181 is considered a promising biomarker for early detection of AD, Frontotemporal lobar degeneration as well as TBI of all severities ^11,23,27-31^. TBI even in its mildest form may have long-lasting consequences in multiple domains of functioning, including cognitive deficits ^32^. While it has been suggested that cognitive tests should be an integral part of the holistic approach of assessment of TBI outcome ^33^, the feasibility of assessing the cognitive impact of minor, non-concussive head impacts have not been widely studied.

To date, no study investigating the effect of repetitive non-concussive head impacts such as heading in soccer on the plasma level of pT181 has been performed. Here we aimed to test the hypothesis that such impacts can lead to quantifiable changes in the plasma level of specific neuroproteins and these changes are accompanied by neurocognitive impairments at multiple time points after the impacts. We focused on quantifying plasma levels of total and phosphorylated tau in highly uniform cohort, young male soccer players, before and after the physical training and regular heading training sessions.

## METHODS

### Participants

The experimental group included soccer players that underwent blood sampling and focused psychological tests prior to training sessions, in order to establish the pre-training (reference) baseline in both domains. The individual characteristics of the participating cohort are summarized in Table 1.

**Table 1.**
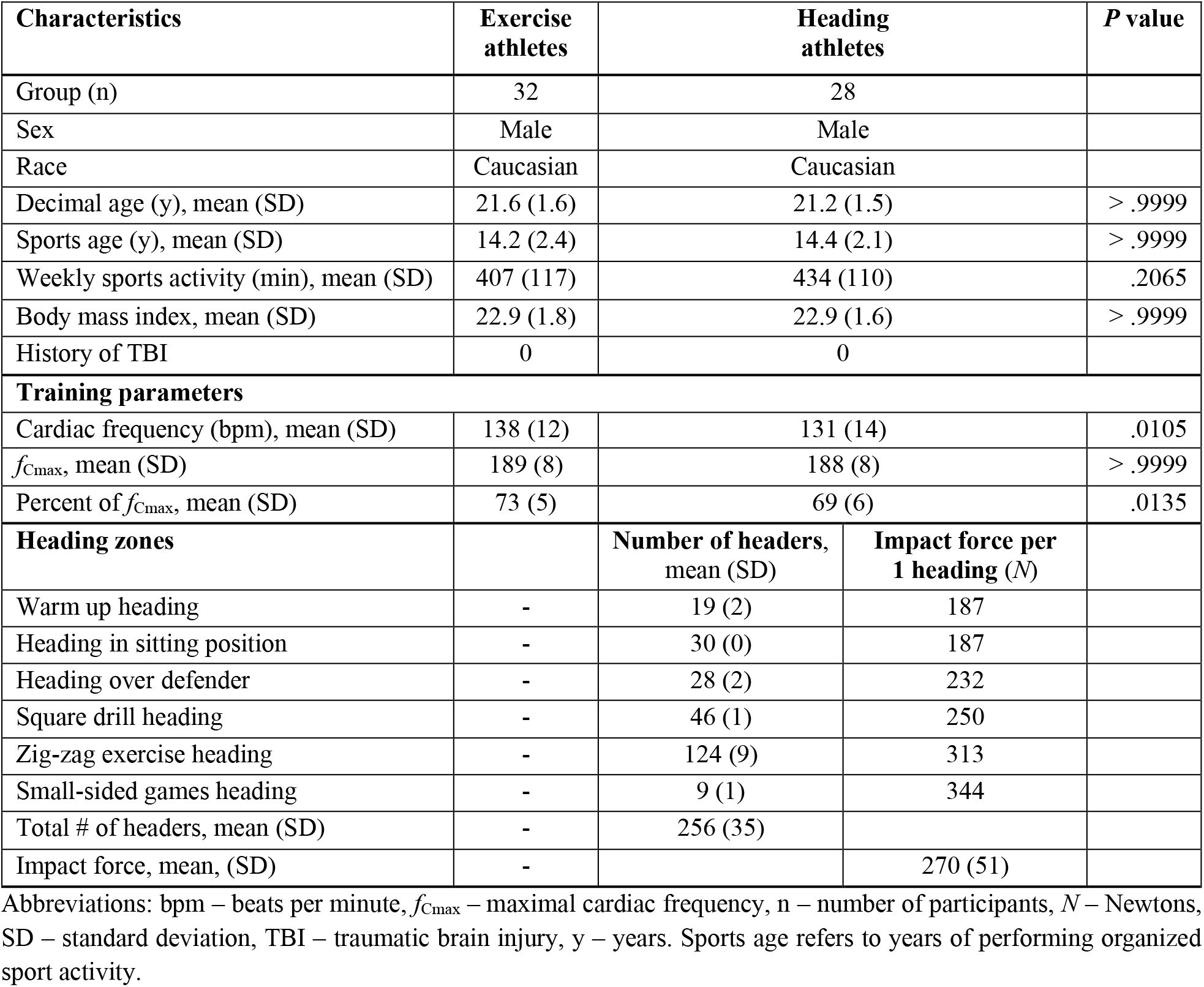
Characteristics of the study participants, training parameters and heading zones.

The Ethical review board of the Comenius University in Bratislava, Faculty of Physical Education and Sport approved this study with human participants and all involved athletes provided written informed consent prior to beginning the study.

### Study design

All athletes were evaluated for physical fitness by recording their individual cardiac frequency (*f*_C_) using a heart rate monitor Polar Team 2 Pro H10 in 5-second intervals (Polar Electro Oy, Kempele, Finland). Individual baseline performance of the athletes and the maximal cardiac frequency (*f*_Cmax_) were measured during a probe training 7 days before the start of the study. The highest *f*_C_ during the test was determined as *f*_Cmax_. The acquired data served as an indicator for designing the consecutive soccer training and heading sessions. This strategy aimed to maintain a high level of physical activity that was within the aerobic interval, represented by 70-75% of *f*_Cmax_. Furthermore, this target load intensity allowed simultaneous performance of aerobic soccer exercise and successful accomplishment of header tasks during the heading training. Except of the training units, the players followed a resting regime for 24 hours before and between blood collection time points. Probe training (week 0) was followed by soccer training without heading the ball (week 1) and subsequent soccer training including the ball heading drills (week 2).

### Soccer Training protocol

All trainings in the experiment were strictly planned and controlled by professional trainers to achieve comparability to the highest possible extent. The soccer training unit (hereafter referred to as exercise, abbreviated as “E”) comprised a warm-up (22 min), two exercise zones (58 min), and a final compensatory exercise (5 min) with a total duration of 85 min. The warm-up zone included running (7 min), followed by exercise without the ball (6 min) and exercise with the ball (9 min). The main part of the training unit consisted of 2 different small-sided games. Players were not allowed to do headers during the entire soccer training unit.

### Heading protocol

Heading training (abbreviated “H”) followed a modified physical exercise protocol when compared to soccer training. All zones, including the warm-up and main training units contained the heading of the ball. The warm-up included two sets of three-meter hand passes of the ball to the head of a teammate. The main heading training included 5 zones of heading exercise: 1. Heading after hands pass in a sitting position (sit with the spread legs) for three sets, each set with 10 attempts of heading; 2. Heading over the semi-active defender after hands pass in two sets of 15 attempts each; 3. Heading in the square for four sets, each set lasted 3 minutes followed by 30-second rest interval; 4. Heading in the zig-zag exercises for four sets, each set lasted 3 minutes with 30-second rest interval; 5. Five small-sided games (SSG) with a given pitch size (25×18 meters) and a rule that a goal can only be scored with a header. During the SSG, hand pass and a head pass were continuously repeated. Each small-sided game lasted 2 minutes followed by 2-minute rest interval. The total duration of the heading training was 85 min.

### Impact force

The average impact force (*F*) for specific heading zones was calculated using the formula:

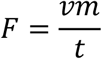

Assuming inputs (*v*) as incoming velocity (m/s), (*m*) mass of the ball (kg) and (*t*) impact duration (s) ^34^.

Incoming velocity of the ball in specific zones during heading exercise was quantified using the radar LM600 (UNI-Trend Technology). The measured velocities of the ball were as follows: warm up heading (8.3 m.s^-1^), heading in sitting position (8.3 m.s^-1^), heading over defender (10.3 m.s^-1^), square drill heading (11.1 m.s^-1^), zig-zag exercise heading (13.9 m.s^-1^), small-sided games heading (15.3 m.s^-1^). The ball mass (0.45 kg) was considered as indicated by the manufacturer. Throughout the study, only officially tested FIFA Quality Pro marked balls (Adidas Uniforia League Pro) were used. The average time of impact duration (0.02 s) was adopted according to published data for ball size 5 and inflation pressure of 15 psi (1 bar) at the speed of 15 m.s^-1 35^ considering the velocity as a most important factor contributing to the peak impact force ^36^.

All the ball parameters listed are in accordance with the International Football Association Board 2020/21 Laws of the game, Law 2 ^37^.

The ball heading training represents a pool of low-intensity head impacts that correspond to the practice of heading techniques routinely performed in youth to adult categories in soccer. The average impact force of 270 *N* as quantified in this study is substantially lower compared to peak impact forces of soccer balls (1700 *N*) measured at higher speeds in the elite soccer league ^35,38,39^. High-intensity head impacts, which are typically played during corner kicks, goalkeeper kickouts, or long-distance passes, were not performed in this study.

### Video analysis

Athletes were monitored by a video system during both training units. Video analysis was used to cross-check the number of ball headings for each player in specific training zones.

### Blood collection and processing

The blood collection was performed by professional medical personnel at 3 time points: before training, 1 h, and 24 h after each training session. Venous blood was collected in a nonfasting state in K2EDTA tubes (Greiner Bio-One, Austria) and placed on ice until processed. All blood samples were centrifuged within 60 minutes from the time of blood draw, at 2000 x *g* for 10 minutes at 4°C, and plasma was isolated. Prepared aliquots were stored at −80°C until assayed.

### Biochemical procedures

Total tau concentrations in plasma were measured by digital ELISA using the Simoa Tau Advantage Kit (Quanterix, Cat.No. 101552). Tau protein phosphorylated at Thr181 (pT181) was quantified using the Simoa pTau-181 V2 Advantage Kit (Quanterix, Cat. No. 103714). The samples were blinded prior to analysis and all assays were measured on a HD-X Analyzer (Quanterix Corp, MA, USA). The average coefficients of variation of measurement of Tau and pT181 were 3% and 4%, respectively.

### Psychological testing

To monitor cognitive changes after soccer and heading training the Trail Making Test (TMT) was used ^40^. The testing consisted of two parts: part A is focused on intensity and sustaining of attention; part B is focused on cognitive flexibility. All participants were tested before and after completing the training units, at timepoints aligned with the blood sample collection. In the Trail Making Test A, a subject was instructed to connect a set of 25 numbered dots in ascending order (1-2-3-4 etc.) as quickly as possible while maintaining accuracy. In a Trail Making Test B, a subject was instructed to connect the dots in sequence while alternating numbers and letters (1-A-2-B-3-C etc.). Time needed to complete the test was recorded and the results are displayed in seconds. Shorter time represents a better score.

### Statistical analysis

Statistical analyses and figure processing were performed using GraphPad Prism (v. 9.3.1) (GraphPad Software, La Jolla, USA). To correct for the basal, interindividual variability among the athletes, the plasma proteins concentration for each individual is expressed as a fold change at 1 h and 24 h from its own baseline. Group changes were visualized as mean fold change profiles with 95% confidence intervals (CI) per training arm. The mean fold change from baseline was compared in both training arms and at all time points (baseline – 0 h, 1 h and 24 h) using ANOVA, with Tukey’s post hoc test. The adjusted *P* < .05 values for multiple comparisons are considered significant. Asterisks indicate statistically significant change from baseline at individual time points (\**P* < .05, \*\**P* < .01, \*\*\**P* < .001).

## RESULTS

### Participants and training parameters

The study included male college soccer players who did not differ significantly in demographic variables, such as age, years of performing organized sport, weekly sport activity, body mass index, and history of TBI (Table 1). Physical performance of all athletes was monitored by heart rate monitors during both training sessions. Comparison of heart rate parameters revealed significantly higher cardiac frequency of soccer athletes during the exercise without heading the ball when compared to the heading training (*f*_C_:138 bpm *vs*. 131 bpm, *P* = .0105). Both types of training also differed in the mean percentage of maximal cardiac frequency that was higher during exercise without headers compared to heading training (*f*_Cmax_: 73% *vs*. 69%, *P* = .0135), indicating more intensive physical exercise (Fig. 1A-B). However, it should be noted that both types of soccer training are within the same aerobic interval of physical performance, representing physiologically comparable groups. Nevertheless, this small difference in physical performance can be due to the higher level of concentration of athletes on the successful completion of ball heading tasks, which resulted in a slightly lower level of physical activity in this specific group.

**Fig. 1.**
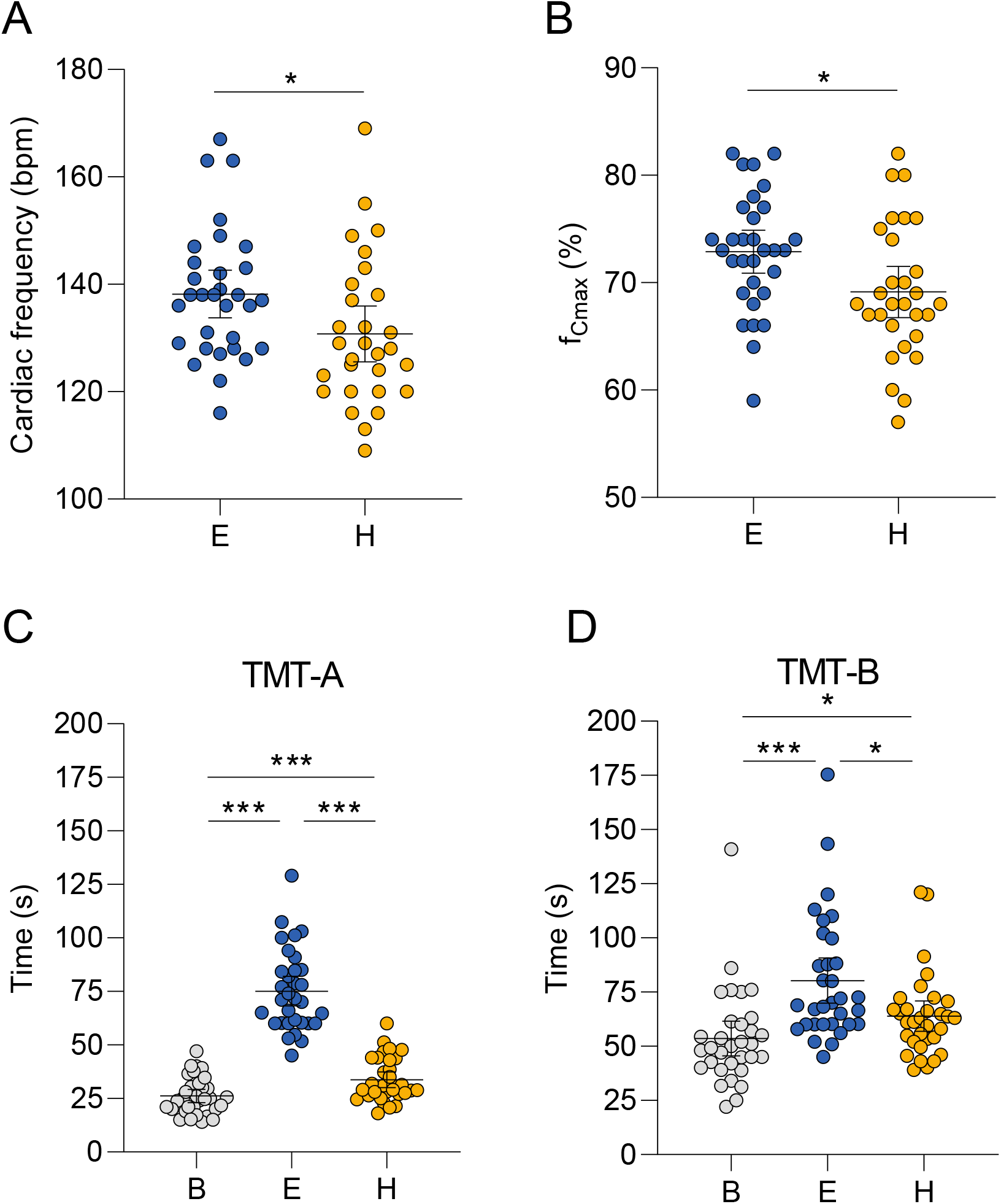
Physical activity parameters and psychological testing. Heart rate monitoring during training units revealed that athletes performed soccer training with higher physical activity compared to heading training, represented by the higher average cardiac frequency (A) and higher percentage of maximal cardiac frequency - f_Cmax_ (B). Soccer exercise and heading training negatively affect the intensity and sustaining of attention of athletes when compared to baseline as revealed by the Trail Making Test A (C). Both training sessions also showed negative outcome on cognitive flexibility of athletes (TMT-B) compared to baseline (D). Psychological testing revealed that non-heading exercise had a more negative impact than heading training. Horizontal black lines denote the mean and 95% CI. B - baseline, E – soccer exercise without heading the ball, H – soccer exercise including heading the ball.

### Cognitive outcomes

Neuropsychological examination of soccer players using both parts of the TMT identified significantly worse scores after exercise and heading training compared to baseline testing (Table 2). Both trainings negatively affected focused attention and cognitive flexibility; however, exercise training without headers had a more negative impact than heading training (Fig. 1C-D).

**Table 2.**
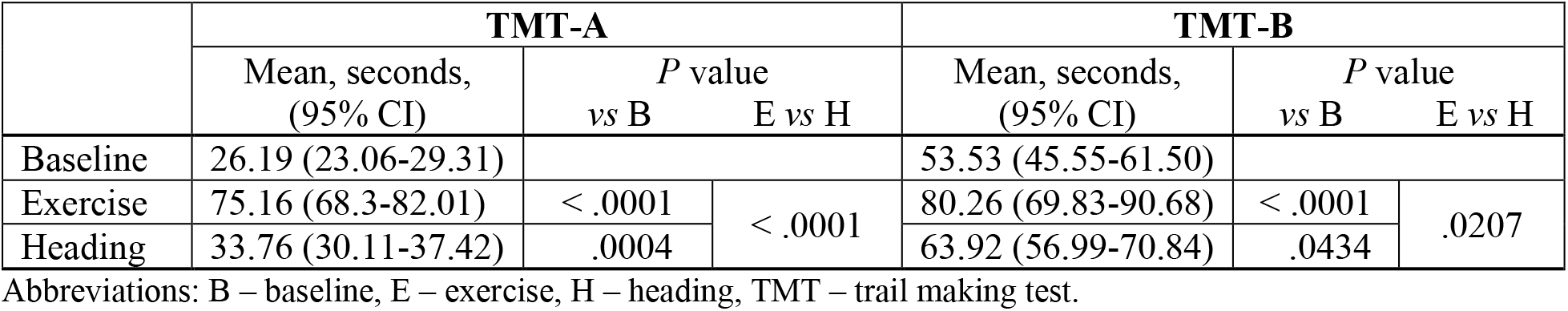
Changes in attention and cognition after exercise and heading training.

### Changes in circulating plasma neuroproteins after exercise and heading training in soccer players

To elucidate the effect of repetitive non-concussive head impacts, we performed a detailed analysis of total tau and phosphorylated tau (pT181) in plasma of soccer players while accounting for the effect of physical activity.

The mean plasma concentrations of total tau and pT181 examined at baseline were similar in both groups of athletes (Fig. 2A). Quantitative digital ELISA identified significant change in plasma tau levels 1 h after the soccer exercise (1.35-fold, 95% CI 1.2-1.5, *P* = .0001), and 1-hour post-heading training (1.3-fold, 95% CI 1.2-1.4, *P* < .0001) when compared to baseline. The increased total tau levels in plasma of soccer players normalized to baseline within 24 hours (Fig. 2B). Identical profile, yet with a slightly higher magnitude of change, was observed for pT181 at 1-hour post-exercise (1.4-fold, 95% CI 1.3-1.5, *P* < .0001) and post-heading (1.5-fold, 95% CI 1.4-1.7, *P* < .0001), respectively. An increased level of peripheral phosphorylated tau was identified even 24 hours post-training (Fig. 2C). We - observed an increased plasma pT181/Tau ratio in both groups at 1-hour post-exercise (1.1-fold, 95% CI 1.0-1.2, *P* = .0362) and post-heading (1.2-fold, 95% CI 1.1-1.2, *P* < .0001). However, the pT181/Tau ratio remained significantly higher in heading group (1.2-fold, 95% CI 1.1-1.3, *P* = .0024) when compared to baseline even after 24 hours. In contrary, the athletes performing soccer exercise without heading the ball showed a small decreasing trend of pT181/Tau ratio (1.08-fold, 95% CI 1.0-1.16, *P* = .0793) to pre-training level within 24 hours (Fig. 2D). These results show that levels of total tau protein were lower in the heading group, while the fraction of phosphorylated tau still remained at the same level in both groups. Therefore, the amount of pT is relatively higher in the heading group.

**Fig. 2.**
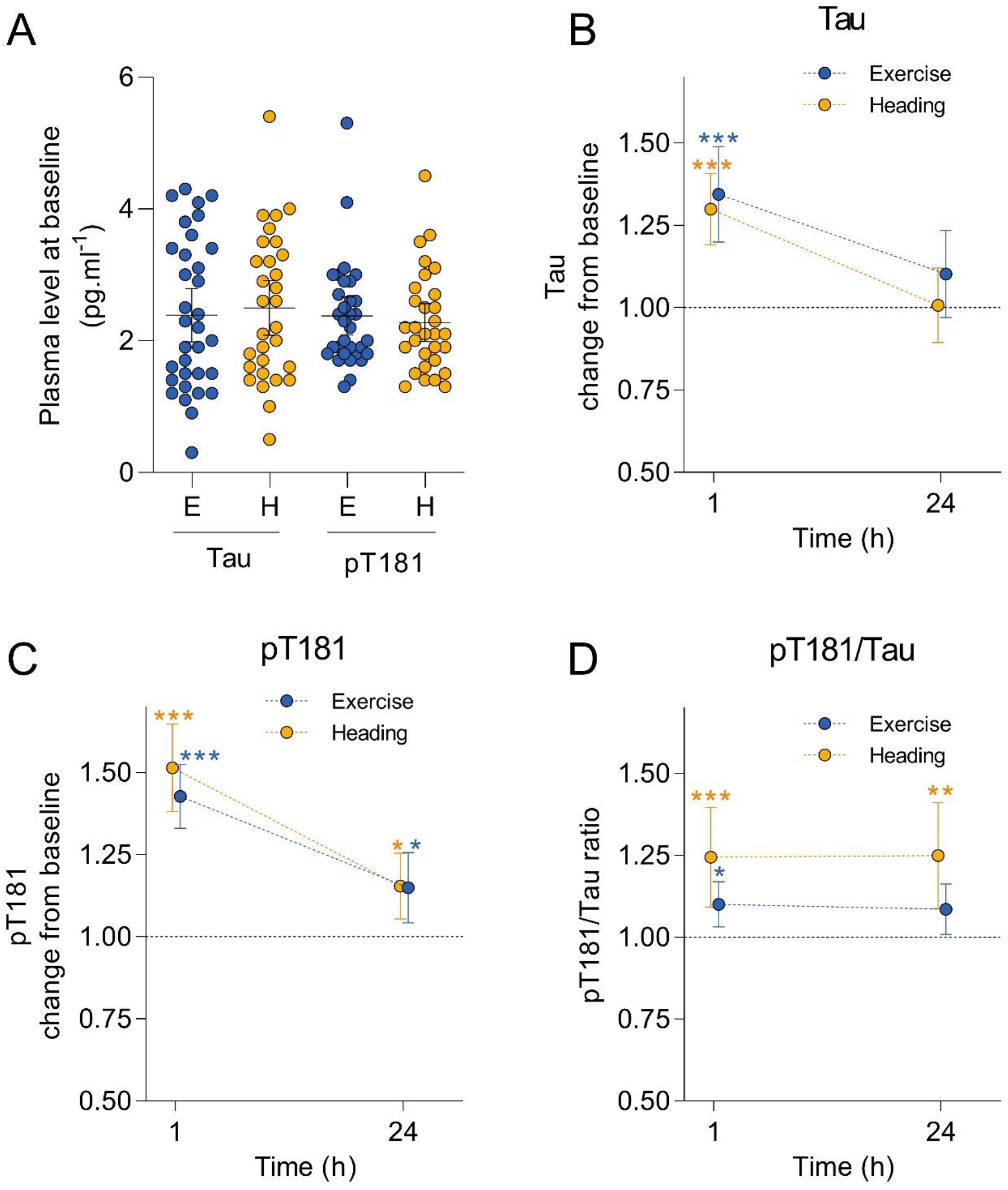
Tau and pT181 Tau levels after soccer exercise and heading training. Analysis of total tau and phosphorylated tau protein at threonine 181 (pT181) in plasma reveals no difference between the exercise – E (blue) and heading – H (orange) groups at baseline (A). Horizontal black lines denote the mean and 95% CI. Significant change in plasma tau from baseline was observed in athletes 1 hour after exercise and heading training. The elevated tau levels normalized to baseline (black dotted line) within the 24 hours (B). Significantly increased plasma pT181 levels were observed 1 h after both types of training, and elevated levels were still detected after 24 hours (C). pT181/Tau ratio was significantly higher 1 hour after exercise and heading, and remained specifically elevated in the heading group even after 24 hours (D).

## DISCUSSION

The main outcome of our study is the observation that physical exercise as well as the ball heading in soccer cause a highly significant increase in the population of tau proteins phosphorylated at Threonine 181, which is a promising marker of neurodegeneration. The fraction of total tau protein was found increased in control athletes and athletes after concussion and TBI in several studies ^41,42^. Here, we provide direct evidence that tau protein level is increased in the blood as a consequence of physical training, which is consistent with our previous observations ^43^. However, the significant increase in the level of phosphorylated tau protein (pT181) in the plasma of soccer players after regular physical soccer training and non-concussive repetitive heading of the ball, which is a usual practice in the game, is surprising. Such increase has not been previously reported in the literature. The finding was not expected in the exercise cohort, since the phosphorylation of tau protein is typically associated with TBI and moreover, it is one of the early diagnostic and prognostic biomarker candidates of AD dementia ^10,11,27,28^. However, it should be noted that the 1.5-fold increase of pTau after mild heading training in this study was lower compared to the increase observed in mild or severe TBI cases ^28,42^. Increased pT181 even after 24 hours indicates longer persistence of phosphorylated tau in the brain. Based on the data, we emphasize the increase of tau and pT181 tau after physical training as an important contributing factor to consider when evaluating tau levels in individuals suffering from sport-related head traumas. Furthermore, the results of our study provide valuable information to determine the analytical threshold for measuring the pT181 tau level in future TBI research and clinical practice.

Interestingly, the pT181/Tau ratio remains significantly higher specifically after mild repetitive head impacts. This also suggests that the population of tau proteins is enriched by phosphorylated tau fraction, which may have long-lasting consequences in the brain of head-impacted individuals. The increased amount of pTau molecules may serve as substrate for accumulation of pathological forms of tau after sufficient period of time.

In the light of the recently published findings ^28^, the repetitive head impacts may lead to the pathological changes in long-term perspective. We hypothesize that even the slight increase in the ratio of pT181/Tau, which we have observed after the non-concussive repetitive head impacts, may represent a major difference for the individuals playing contact sports versus other head impact-free physical activities. It cannot be excluded that even non-concussive, very mild, multiple head impacts can lead to the development of neurocognitive disorders later in life. Contact sports may therefore impose a certain degree of health risk promoting the silent development of specific tauopathies (CTE, AD, etc.) and spreading of the pathology, as was indicated in the recent epidemiological study ^7,44^. Since the upregulation of phosphorylated Tau could precede the development of the obvious pathological signs in behavior ^30^, we urge more precautions in contact sports, especially those in which, although very mild, repetitive head impacts are frequent such as ball heading in soccer. It is especially important for at-risk groups, such as young adults in the development, and athletes with longer total exposure time.

Another surprising result of our study was finding that non-heading exercise had a more negative impact on cognition than heading training; we expected a more intense deterioration of cognitive parameters in the heading group. Assuming the higher physical performance in the exercise group, we hypothesize that even slightly increased physical activity was sufficient to induce the higher mental fatigue of athletes, which negatively affected the recorded cognitive parameters – the focused attention and cognitive flexibility. The data are consistent with published evidence of decreased psychophysiological and cognitive responses related to mental fatigue after soccer training ^45,46^. This effect was also observed in elite young basketball players ^47^ and table tennis players ^48^. Concerning to the possible link between tau protein, intensity of physical activity and cognition, the positive effect of physical activity in non-contact sports was associated with slower rate of cognitive decline regardless of tau protein levels in a longitudinal perspective ^49^. In our study focused on acute effects we did not observe any correlation between tau protein levels and cognitive parameters as well.

### Strengths and limitations

The design of the study accounted for the factor of physical activity in studying the effect of mild repetitive head impacts. Our experimental groups were relatively uniform, comprising athletes who did not differ significantly in demographic variables. In addition to the quantified parameters of physical activity, we also determined the number and intensity of head impacts, which increases the reproducibility of the study results.

Nevertheless, we acknowledge several limiting aspects of the study that need to be considered. In particular, we determined the level of phospho-fraction of tau protein by measuring only one phosphorylated epitope, threonine 181. More complex data should be collected e.g. quantifying other relevant tau populations with different phosphorylated epitopes such as T217 and T231 and others. Consequently, it remains to be elucidated whether the increased level of pT181 is pathological or physiological. Another limitation of the study is the number of participants, although this is a relatively very uniform cohort; the results should be validated in a larger cohort using additional follow-up time points. As this cannot be done by one centre/group, our study thus can initiate many of these studies in the near future. It will be also important to determine whether the same findings are confirmed in female athletes. Based on our results, we acknowledge that experimental group of mild repetitive headers without any physical activity could help to further elucidate the complexity of physiological effects observed in this particular study.

## CONCLUSION

We identified elevated levels of pT181 tau, a biomarker of AD and TBI, as well as an increase in pT181/Tau ratio, in the plasma of subjects after physical exercise alone and after repetitive non-concussive head impacts. The ratio pT181/Tau, which remained elevated in the head impact cohort specifically, may indicate a prolonged molecular life span of phosphorylated tau protein in the brain, which may serve as a substrate for accumulation of pathological tau protein structures leading to neurotoxicity later in life.

## Data Availability

All data produced in the present study are available upon reasonable request to the authors

## Author contribution

Study design, JP, PP, IT, MM, MS, MC, PF; investigation, JP, PP, IT, JH, SP, MS, BK; data curation, JP, MC, MS, BK; writing–original draft preparation, MC, PF; writing–review and editing, IJ, MM, JP; funding acquisition, MM, PF, MC, JP, PP. All authors have read and approved the final version of the manuscript.

## Funding

This research was co-funded by the Slovak Research and Development Agency, grant number APVV-17-0668, APVV-19-0568, APVV-20-0615 and the Ministry of Education, Science, Research and Sport of the Slovak Republic, in the form of research grants VEGA 2/0118/19, 2/0153/22 and ERA-NET Neuron Neu-Vasc. The funding source had no role in study design, data collection and analysis, decision to publish, or manuscript preparation.

## Conflict of interest

The authors declare no conflict of interest.

## Acknowledgements

We would like to thank the study participants and collaborators including Silvia Putekova, Jana Martinkova for their valuable help in obtaining the blood samples. We thank Denisa Hazerova, Phuong Truc Pham, Samuel Paulik and Patrik Sivco for their help in implementing the cognitive tests.

## Notes

### Competing Interest Statement

The authors have declared no competing interest.

### Funding Statement

This study was funded by by the Slovak Research and Development Agency, grant numbers APVV-17-0668, APVV-19-0568, APVV-20-0615 and the Ministry of Education, Science, Research and Sport of the Slovak Republic, in the form of research grants VEGA 2/0118/19, 2/0153/22 and ERA-NET Neuron Neu-Vasc.

